# Lifespan EEG reference charts disentangle alpha-band alterations in neurological and neuropsychiatric disorders

**DOI:** 10.64898/2026.09.19.26363457

**Authors:** Vardhan Paliwal, Alexander Moiseev, Sam M. Doesburg, Pengcheng Xi, Joel S. Winston, Mark P. Richardson, Roman Rodionov, Urs Ribary, Andrew Blaber, George Medvedev, Vasily A. Vakorin

## Abstract

Accurate quantification of alpha-band (8-13 Hz) slowing, a putative biomarker in neurological and neuropsychiatric disorders, is hindered by two problems: the conflation of oscillatory activity with aperiodic 1/f components in EEG spectra, and the absence of age-specific normative references in routine clinical EEG. Both problems are compounded by pronounced changes in alpha rhythms across the lifespan. Here we generate lifespan brain reference charts for alpha peak frequency and power, deriving lobe-specific models from raw and 1/f-adjusted spectra in a large outpatient cohort (N = 22,094; age: 1-100 years). We then applied these normative models to inpatients spanning a range of mental disorders. Across all diagnostic groups, alpha power was consistently reduced relative to age-matched norms, regardless of whether spectra were 1/f-adjusted. In contrast, alpha peak frequency showed a striking method-dependent dissociation: conventional analysis of raw spectra suggested alpha slowing, whereas 1/f-adjusted estimates revealed robust alpha acceleration. These findings demonstrate that aperiodic components can mask underlying oscillatory changes, and that 1/f adjustment is essential for interpreting alpha dynamics in clinical populations. More broadly, these findings indicate that the widely reported “alpha slowing” in mental disorders may partly reflect unmodeled shifts in aperiodic neural activity rather than a true deceleration of alpha oscillations.

## 1. Introduction

Alpha-band (8-13 Hz) rhythms are the dominant oscillatory feature of human electroencephalograms (EEG) and play a crucial role in cognition [1], exhibiting sensitivity to both transient physiological states (e.g., eyes open vs. closed) and stable inter-individual constitutional differences. This natural variability makes alpha rhythms a valuable biomarker accessible during routine clinical EEG assessment, where alpha frequency, symmetry, and reactivity are systematically evaluated and guide diagnosis and prognosis of encephalopathy, coma, dementia subtyping, and developmental maturation across neurology practice [2]. For instance, alterations in alpha rhythm parameters (peak power and frequency) follow a consistent pattern across a wide range of neurological and neuropsychiatric conditions: an apparent alpha slowing, reflected in reduced peak frequency [3–8], accompanied by reduced alpha power [3–9]. However, determining what constitutes the ‘slowing’ or ‘reduced power’ in an individual patient remains challenging. Alpha parameters exhibit substantial inter-individual variability, influenced by age, sex, and other factors, and undergo systematic age-related changes across the lifespan. Without normative reference charts explicitly modeling this variability, distinguishing pathological deviations from normal age-related trajectories remains challenging [10].

Conventional metrics of alpha activity are typically derived by band-pass filtering or integrating spectral power within a fixed alpha range. These measures show robust age-related trends: both alpha power and peak frequency increase throughout childhood and adolescence [11–13], then decline from early adulthood onwards [4, 5, 12, 13]. Such trajectories are widely interpreted as reflecting maturation and subsequent degeneration of thalamo–cortical and cortico–cortical networks that generate alpha rhythms. However, these conventional estimates are computed on the raw power spectrum and therefore conflate true oscillatory activity with broadband, non-oscillatory components.

A key contributor to this broadband background is aperiodic 1/f activity, which describes the characteristic monotonic decrease of EEG power with increasing frequency [14, 15]. Although previously dismissed as background noise, this aperiodic component comprises a significant portion of cortical electrical activity [16], is sensitive to shifts excitatory-inhibitory balance [17], and is dynamically modulated during cognitive tasks and arousal states [16, 18]. This aperiodic activity is characterized by its slope (the steepness of spectral decay) and offset (a broadband vertical shift in power), both of which vary across the lifespan and in mental disorders [19–24]. Since aperiodic activity contributes to overall alpha-band power, changes in its slope or offset can distort both alpha power and frequency [14]. As a result, conventional alpha metrics may mischaracterize the true oscillatory dynamics they are intended to capture.

Given that aperiodic activity contributes to the overall power in the alpha-band and itself changes across many mental disorders, a central question emerges: do conventional alpha metrics, which conflate oscillatory and aperiodic components, accurately characterize the underlying oscillatory changes? If aperiodic activity is systematically altered in clinical populations with mental disorders, conventional estimates of “alpha slowing” and “reduced power” may partly, or even predominantly, reflect these broadband changes rather than true alterations in alpha oscillations. Yet, to date, no work has systematically contrasted conventional versus 1/f-adjusted alpha parameters in the characterization of mental disorders across the entire lifespan.

We addressed this gap by combining spectral parameterization with normative modeling of clinical EEG. Using a validated spectral parametrization approach [14], we decomposed resting EEG power spectra into periodic alpha oscillations and aperiodic 1/f activity, yielding four parameters: 1/f-adjusted alpha peak frequency and power, and aperiodic slope and offset, alongside the conventional estimates of alpha oscillations: unadjusted alpha frequency and absolute alpha power. We constructed sex- and brain region-stratified normative reference charts for all parameters using a large outpatient EEG cohort (n=22,094), drawn from public hospitals in British Columbia, Canada, spanning ages 1-100 years. To test how mental disorders deviate from these normative trajectories, we applied the same pipeline to inpatient EEGs with linked diagnostic codes from electronic health records (Table 1). For each inpatient, we computed age-matched deviation scores for all spectral parameters of interest (Fig. 1), comparing conventional and 1/f-adjusted alpha metrics, as well as aperiodic features, within and across diagnostic groups. We hypothesized that this multi-parameter approach would reveal nuanced patterns: for instance, deviations in 1/f-adjusted alpha but not in conventional alpha.

**Table 1:** Demographics of Inpatients’ Cohort. Inpatients underwent clinical assessment including symptom evaluation and EEG analysis, and were classified into Case Mixed Groups (CMGs) as defined by the Canadian Institute of Health Informatics (CIHI) and assigned comorbidity scores (0-4) reflecting overall health status. Seven groups of neurological and neuropsychiatric disorders within the CMG category representing mental disorders were selected based on largest available sample sizes, with a minimum threshold of 50 participants for either sex per clinical group.

| Mental Disorder | Sex | Age Range (years) | Number of Inpatients | Mean Age ± SD (years) |
| --- | --- | --- | --- | --- |
| Seizure Disorder | Male | 1 - 93 | 550 | 49.4 ± 26.2 |
| | Female | 1 - 97 | 454 | 53.5 ± 26.5 |
| Status Epilepticus | Male | 1 - 94 | 157 | 52.2 ± 21.3 |
| | Female | 1 - 90 | 152 | 52.4 ± 16.9 |
| Organic Mental Disorder | Male | 20 - 95 | 186 | 71.8 ± 14.2 |
| | Female | 33 - 104 | 136 | 76.7 ± 13.4 |
| Dementia | Male | 40 - 95 | 88 | 75.2 ± 10.9 |
| | Female | 43 - 97 | 59 | 77.3 ± 11.3 |
| Substance Abuse | Male | 20 - 76 | 68 | 49.8 ± 14.3 |
| | Female | 16 - 77 | 27 | 47.4 ± 16.7 |
| Schizophrenia | Male | 17 - 73 | 76 | 37.3 ± 15.9 |
| | Female | 16 - 80 | 55 | 38.2 ± 17.7 |
| Schizotypal Disorder | Male | 11 - 82 | 103 | 35.0 ± 17.2 |
| | Female | 14 - 84 | 108 | 42.3 ± 18.1 |

**Figure 1:**
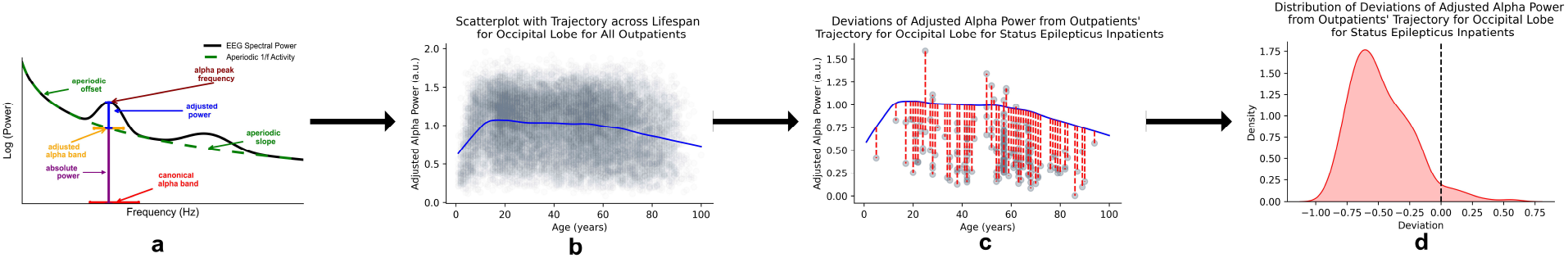
Analysis workflow shown for adjusted alpha power for the occipital lobe and status epilepticus group. **(a)** Spectral parameter decomposition method provided alpha parameters (adjusted alpha peak frequency and adjusted alpha power) after accounting for aperiodic 1/f activity (slope and offset.) **(b)** Lifespan trajectory for adjusted alpha power for occipital lobe for outpatients constructed using scatterplot averaging on the distribution of adjusted alpha power. This serves as the reference trajectory (normative age-related changes for the specific population). **(c)** Adjusted alpha power for status epilepticus patients quantified by projecting them onto the lifespan trajectory and calculating individual deviation scores. **(d)** Distribution of deviation scores visualized and compared against the reference mean of 0 (dotted line). The distribution is right-skewed, implying that the mean deviation in adjusted alpha power in the occipital lobe for status epilepticus inpatients is negative, indicating reduced adjusted alpha power compared to the reference trajectory.

This study had three aims. First, to characterize normative lifespan trajectories for six EEG spectral parameters: conventional alpha frequency and power, 1/f-adjusted alpha frequency and power, and aperiodic slope and offset. Second, to quantify how major mental disorder groups deviate from these normative trajectories across all six parameters. Third, to test whether conventional and 1/f-adjusted alpha parameters show divergent patterns in mental disorders, and whether aperiodic parameter changes explain any observed divergence.

## Results

### Alpha-band power and frequency exhibit characteristic inverted-U shaped trajectories across the lifespan, regardless of the 1/f-adjustment

We first constructed normative brain charts of alpha-band power and peak frequency to quantify their age-related trajectories and assess how these are affected by aperiodic 1/f activity. In outpatient EEGs, we estimated source-level activity, parcellated cortex using the Destrieux atlas [25], extracted spectral parameters for each parcel, and averaged them across six cortical lobes (insular, limbic, frontal, occipital, parietal and temporal). For each region and sex, we then modeled lifespan trajectories for unadjusted alpha power (maximum absolute power within 8–13 Hz), 1/f-adjusted alpha power [14], and their corresponding peak frequencies using a scatterplot-averaging approach (Figs. 2, 3; see Methods 1).

**Figure 2:**
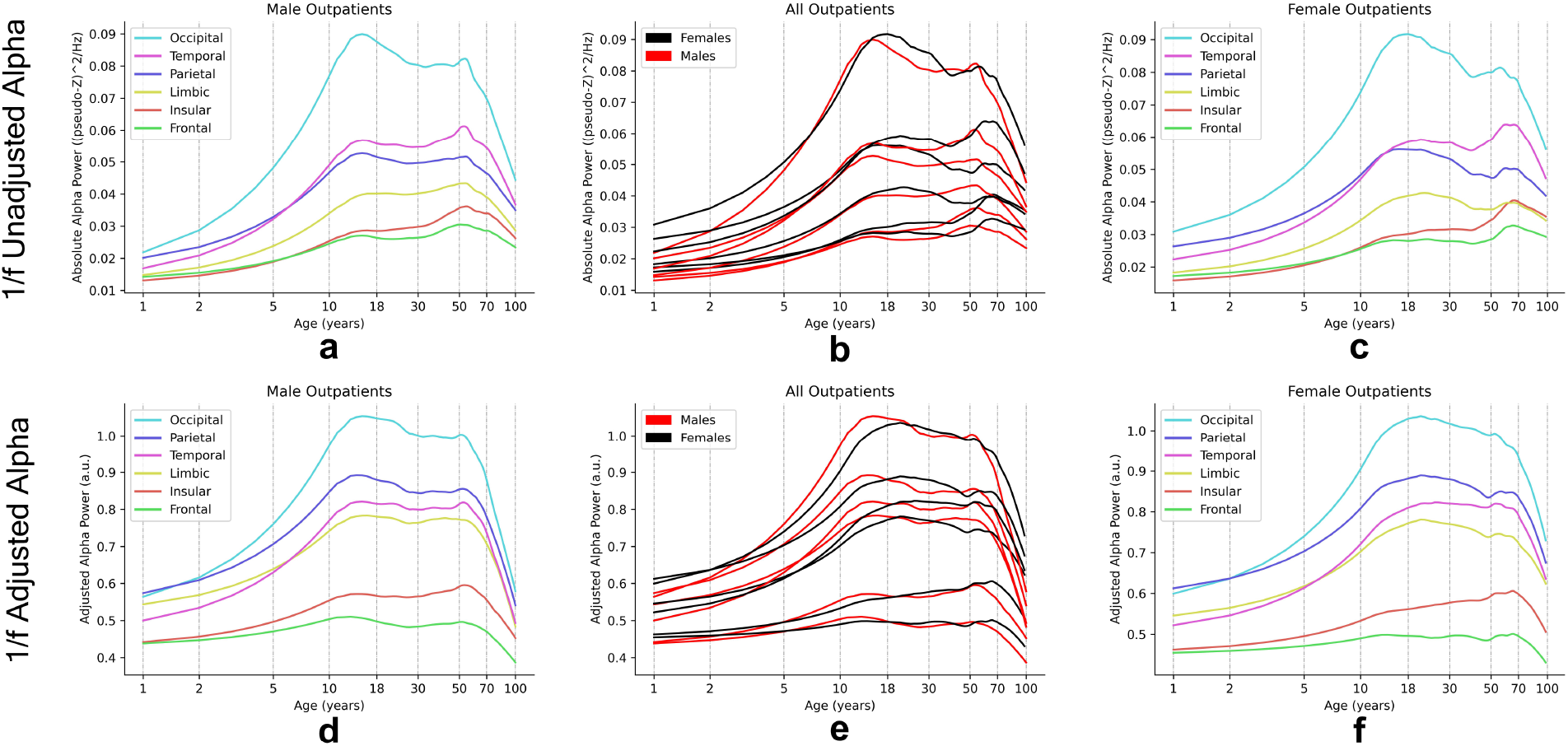
Alterations in alpha power as a function of age across lifespan due to variation in three factors: estimation method, brain region, and sex. The age on the x-axis is represented using natural logarithmic scale. Alpha power across both the estimation methods revealed a stable developmental trend across lifespan: increasing from childhood through adolescence, plateauing in adulthood, and declining in later stages of life. Both estimation methods showed high regional differences. **(a)** Lifespan trajectories for absolute alpha power for males. **(b)** Lifespan reference charts for absolute alpha power for males and females compared together visually. Variability in alpha power between sex across brain lobes is low. **(c)** Lifespan reference charts for absolute alpha power for females. **(d)** Lifespan reference charts for adjusted alpha power for males. **(e)** Lifespan reference charts for adjusted alpha power for males and females compared together visually. Variability in alpha power between sex across brain lobes is low, similar to the absolute alpha power. **(f)** Lifespan reference charts for adjusted alpha power for females.

**Figure 3:**
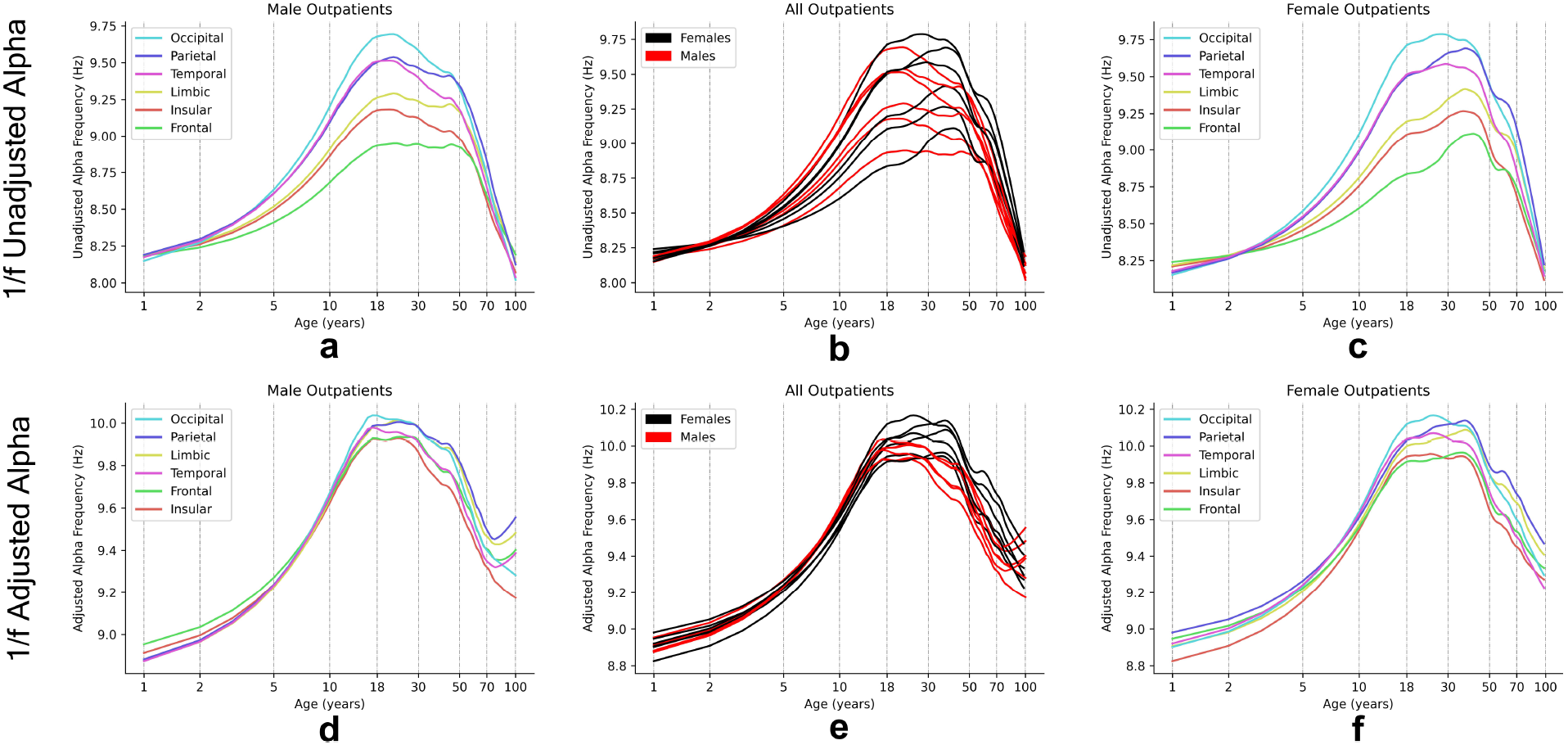
Alterations in alpha frequency as a function of age across lifespan due to variation in three factors: estimation method, brain region, and sex. The age on the x-axis is represented using natural logarithmic scale. Both unadjusted and adjusted alpha frequencies showed inverted U-shaped lifespan trajectories, characterized by an increase through childhood and adolescence, a peak in early adulthood, followed by a plateau through middle adulthood, and a decline in later years. Regional differences were higher compared to sex-differences. Visually, the variability across brain regions was higher at unadjusted alpha frequency than at adjusted alpha frequency. **(a)** Lifespan reference charts for unadjusted alpha frequency for males. **(b)** Lifespan reference charts for unadjusted alpha frequency, males and females compared visually. **(c)** Lifespan reference charts for unadjusted alpha frequency for females. **(d)** Lifespan reference charts for adjusted alpha frequency for males. **(e)** Lifespan reference charts for adjusted alpha frequency, males and females compared visually. **(f)** Lifespan reference charts for adjusted alpha frequency for females.

Lifespan trajectories for both absolute and 1/f-adjusted alpha power followed a characteristic inverted U-shape curve. Alpha power increased from childhood into adolescence, reached a plateau in early-to-mid adulthood, and declined in later life (**Fig. 2a,c,d,f** for representative lobe trajectories stratified by sex; **Fig. 2b,e** for lobes across sex compared together). This pattern was robust across estimation methods, sexes and cortical regions. Across the entire age range, occipital cortex showed the highest alpha power, and inter-regional differences were consistently larger than sex differences (Supplementary Fig. 4a,b).

**Figure 4:**
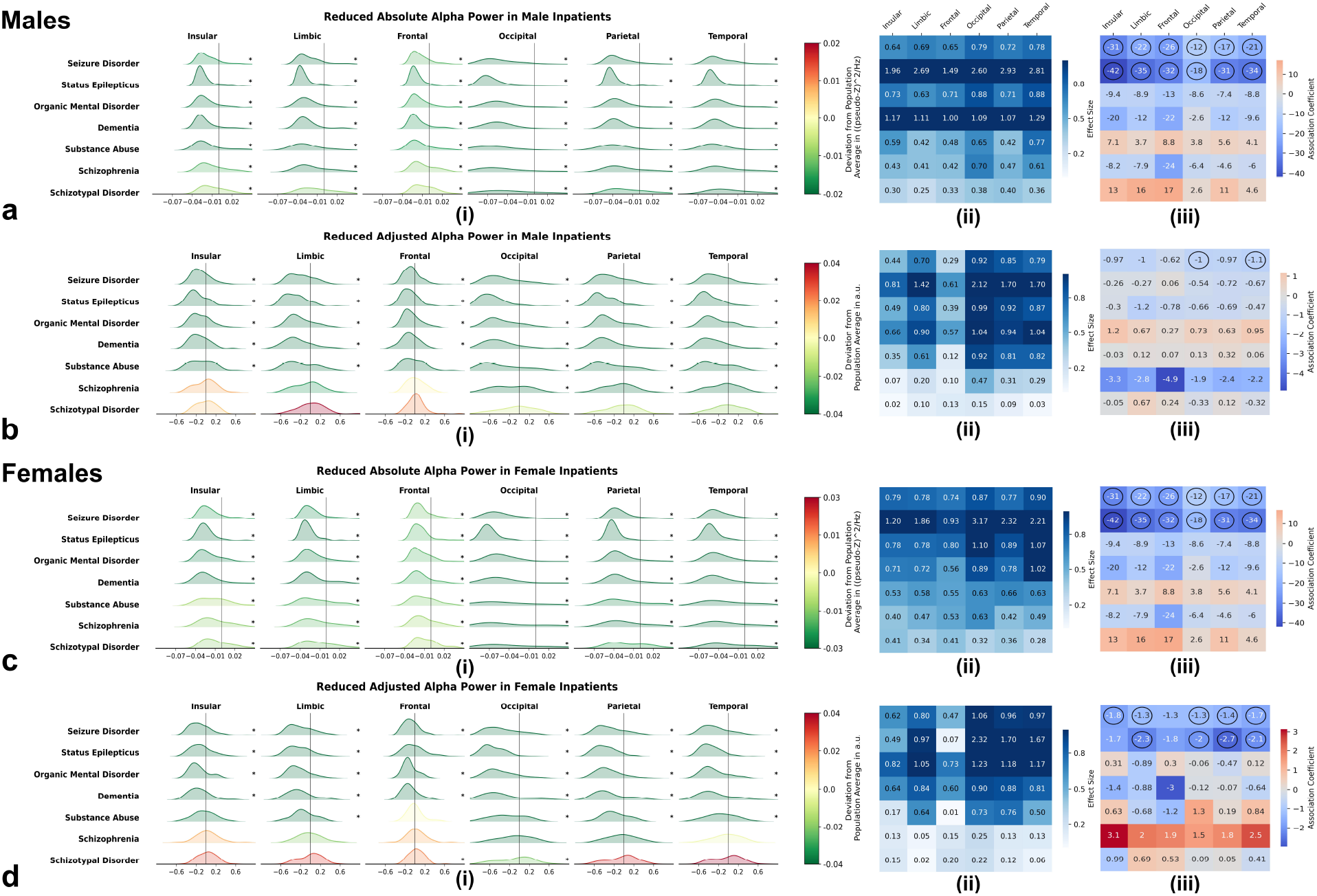
Neurological and neuropsychiatric disorders are characterized by a reduction in alpha power. Panels a-d represent this reduction for different diagnostic groups, stratified by sex, brain region, and estimation method. **a-d(i)** Deviations of inpatients alpha power from normative reference charts. Green distributions represent negative deviations, indicating reduced alpha power, while red distributions represent positive deviations, indicating increased alpha power in clinical groups relative to normative values, with statistical significance marked by (*). All clinical groups showed consistent reductions in both absolute and 1/f-adjusted alpha power. **a-d(ii)** Effect sizes of these reductions. Each cell represents the effect size for a unique disorder-region pair with darker colors indicating stronger effects. The effects were medium to very large, varying across diagnostic groups. **a-d(iii)** Correlations between deviations in alpha power and comorbidity scores, with statistically significant correlations marked by circles. Each cell represents correlation for a unique disorder-region pair. Negative correlations are shown as blue cells and positive correlations in red cells. Seizure disorder and status epilepticus inpatients showed significant negative correlations (*p* < 0.1), implying that with increasing medical burden, these clinical groups showed smaller reductions in both absolute and adjusted alpha powers.

The alpha-band peak frequency (APF) showed a similar global pattern. Both unadjusted and 1/f-adjusted APF increased through childhood and adolescence, peaked in early adulthood, remained relatively stable across middle age and declined in older age (Fig. 3a,c,d,f for representative lobe trajectories stratified by sex; Fig. 5b,e for lobe trajectories across sex compared together). As with alpha power, trajectories were highly conserved across brain regions and between sexes, with regional variation exceeding sex effects (Supplementary Fig. 4c,d).

**Figure 5:**
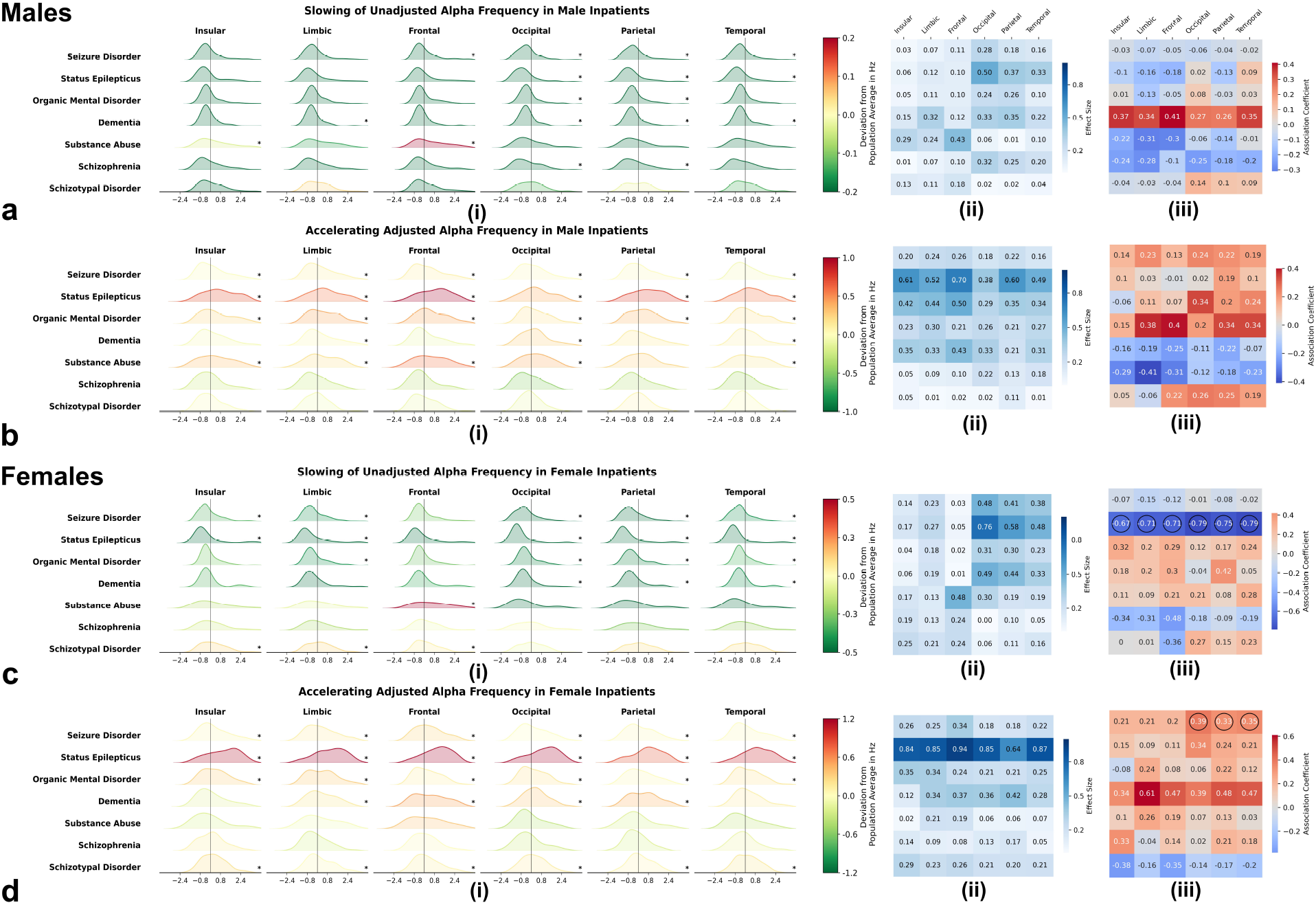
Neurological and neuropsychiatric disorders show alpha acceleration when aperiodic activity is accounted. Panels a-d profiles the changes in alpha frequency, stratified by diagnostic group, sex, brain region, and estimation method. **a**,**c(i)** Deviations in unadjusted alpha frequency from normative reference charts. Green distributions represent negative deviations indicating reduced alpha frequency (alpha slowing) in a wide range of disorders, with statistical significance (*p* < 0.1) marked by (*). This effect was heterogeneous across disorders, sex and brain regions. **a**,**c(ii)** Effect sizes of this slowing, with darker colors indicating stronger effects. The effects were low to medium, varying across diagnostic groups and brain regions. **a**,**c(iii)** Correlations between deviations in unadjusted alpha frequency and comorbidity scores, with statistically significant correlations marked by circles. Negative correlations are shown as blue cells and positive correlations in red cells. Status epileptics showed negative, indicating that unadjusted alpha frequency deviated lesser with increasing medical burden in this group. **b**,**d(i)** Deviations in adjusted alpha frequency from normative reference charts. Red distributions represent positive deviations, indicating increased alpha frequency (alpha acceleration) in neurological and neuropsychiatric disorders, with statistical significance (*p* < 0.1) indicated by (*). This acceleration was relatively stable across brain lobes but showed heterogeneity across brain regions and diagnostic groups. **b**,**d(ii)** Effect sizes of this acceleration. Low to high effect sizes were observed overall, with the effect being greatest in status epilepticus inpatients. **b**,**d(iii)** Correlations between deviations in adjusted alpha frequency and comorbidity scores. Positive, significant correlations (*p* < 0.1) were observed for female status epilepticus inpatients, particularly in the occipital, parietal, and temporal lobes, indicating that greater medical burden is associated with a larger increase in adjusted alpha frequency (higher alpha acceleration) in this group.

Crucially, despite the overall similarities in shape, a key divergence emerged between the two unadjusted and 1/f-adjusted APF in late life. Unadjusted APF showed a steep decline in older adults, with centile values in the oldest individuals approaching those observed in early childhood, creating an almost “circular” lifespan pattern (Fig. 3, Unadjusted APF). By contrast, 1/f-adjusted APF also declined with age but retained clearly adult-like values even in very old individuals and did not regress to infant-like levels (Fig. 3, 1/f Adjusted APF). This dissociation indicates that conventional estimates of “alpha slowing” in ageing are strongly influenced by age-related shifts in aperiodic activity and may substantially overstate actual oscillatory slowing. Together, these findings establish robust, sex-stratified normative reference charts for alpha power and frequency across the lifespan, showing conserved trajectory patterns across cortical lobes for both raw and 1/f-adjusted measures. We next examined deviations from these normative trajectories in clinical populations.

### Alpha power is consistently reduced in neurological and neuropsychiatric disorders, independent of estimation method

Building on our normative lifespan reference charts, we next examined alterations in alpha power across a range of neurological and neuropsychiatric disorders. Because of constraints in diagnostic coding and sample size, we focused on inpatients grouped into broader clinical categories defined by electronic health record classifications (see Methods 1). Within the overarching category of mental disorders, we selected seven subgroups with the largest available samples. Each inpatient was also assigned a comorbidity score that indexes overall medical burden (higher values indicating greater comorbidity). Using the sex- and lobe-specific brain charts derived from the outpatient cohort, we projected absolute and 1/f-adjusted alpha power for each inpatient onto the corresponding normative curves and computed individual deviation scores. Across all diagnostic groups, both absolute and adjusted alpha power were systematically reduced relative to age-matched norms (**Fig. 4 a-d(i)**). This reduction was consistent across estimation methods, sexes, and all six cortical lobes, indicating that reduced alpha power is a robust, method-independent feature of these clinical populations.

To quantify the magnitude and spatial distribution of these deviations, we calculated effect sizes for alpha power reductions within each diagnostic group and cortical lobe. Deviations ranged from medium to very large effect sizes and varied systematically by disorder (**Fig. 4 a-d(ii)**). Seizure disorder and status epilepticus showed the most pronounced reductions, whereas schizophrenia spectrum disorders exhibited comparatively smaller effects. Spatially, occipital, parietal and temporal lobes showed larger mean reductions than insular, limbic and frontal regions. The occipital lobe displayed both the largest effect sizes and the greatest inter-individual variability in deviation scores, whereas frontal regions showed the smallest and least variable effects. Thus, diminished alpha power appears to be a transdiagnostic feature of mental illness, but both diagnostic category and cortical region strongly modulate its severity.

We then examined whether overall medical burden modulates these alpha power deviations by relating individual deviation scores to comorbidity scores. Across diagnostic groups, and particularly in seizure disorder and status epilepticus, higher comorbidity scores were associated with less negative alpha power deviations, that is, values closer to the normative reference (nominal *p* < 0.1; **Fig. 4 a-d(iii)**). Counterintuitively, this negative correlation indicated that patients in poorer overall physical health showed attenuated disorder-related reductions in alpha power. This pattern suggests that general health status may partially offset, or at least obscure, disorder-specific alterations in alpha power in inpatient settings.

In summary, these results demonstrate a consistent and robust reduction in both absolute and 1/f-adjusted alpha power across diverse neurological and neuropsychiatric disorders, most prominently in the posterior cortices and in seizure-related conditions. Reduced alpha power thus emerges as a core neurophysiological alteration in these clinical groups, with its expression shaped by diagnostic category, cortical topography and overall medical comorbidity.

### Alpha oscillations accelerate, not slow, after adjustment for aperiodic activity in neurological and neuropsychiatric disorders

Having established normative lifespan trajectories for alpha-band peak frequency (APF), we next examined how these timing features deviate in neurological and neuropsychiatric disorders, paralleling our analyses of alpha power across diagnostic groups. For both unadjusted and 1/f-adjusted APF, we projected individual values onto the sex- and lobe-specific normative brain charts and computed age-corrected deviation scores across diagnostic group, sex and cortical lobe (Fig. 5).

Unadjusted APF showed a predominant slowing in neurological and neurospsychiatric disorders relative to normative reference values (**Fig. 5 a,c(i)**). However, this slowing was heterogeneous: its magnitude and even its presence varied across diagnostic groups, sexes and brain regions. More consistent slowing was observed in occipital, parietal and temporal lobes, with low-to-medium effect sizes overall (**Fig. 5 a,c(ii)**), and was most pronounced in status epilepticus and dementia. Regionally, occipital cortex showed the strongest slowing, whereas frontal lobes showed the weakest and most variable effects. In contrast, disorders such as substance abuse and schizophrenia spectrum diagnoses exhibited more variable patterns across lobes and sexes, indicating that unadjusted APF slowing is not a uniform signature of mental illness.

In striking contrast, 1/f-adjusted APF generally increased (accelerated) across the same mental disorder groups (**Fig. 5 b,d(i)**). This acceleration was more consistent across cortical lobes than the unadjusted APF slowing, but still showed sex- and diagnosis-specific heterogeneity. Effect sizes ranged from low to high, with the largest accelerations again observed in status epilepticus, particularly in posterior regions (**Fig. 5 b,d(ii)**). Across disorders, occipital, parietal and temporal regions tended to show the most pronounced accelerations, mirroring the regional profile of unadjusted APF slowing but with an opposite direction of effect. Together, these results reveal a clear method-dependent dissociation: when aperiodic activity is not accounted for, APF appears to slow; when it is, APF tends instead to accelerate in mental disorders.

We then asked whether overall medical burden modulates these APF deviations by correlating individual deviation scores with comorbidity. For unadjusted APF, a nominally significant negative correlation with comorbidity emerged in female inpatients with status epilepticus (**Fig. 5 c(iii)**), indicating that higher comorbidity was associated with smaller negative deviations, that is, unadjusted APF closer to normative values. Conversely, for 1/f-adjusted APF, we observed the opposite pattern in female seizure disorder inpatients: deviation scores showed a positive correlation with comorbidity (**Fig. 5 d(iii)**), indicating that higher comorbidity was associated with greater APF acceleration relative to normative reference value. These findings suggest that the impact of general health status on APF is contingent on both the APF metric (unadjusted vs 1/f-adjusted) and the clinical subgroup, potentially reflecting distinct or compensatory pathophysiological processes.

Taken together, these results demonstrate that neurological and neuropsychiatric disorders are characterized by paradoxical signatures in alpha peak frequency: unadjusted APF predominantly slows, whereas 1/f-adjusted APF accelerates. This divergence, and its sensitivity to sex, cortical region and comorbidity, indicates that conventional APF measures and aperiodic-adjusted metrics capture different facets of neurophysiological dysregulation.

### Lifespan trajectories show progressive flattening and attenuation of aperiodic 1/f activity

To characterize age-related changes in background neural activity and thereby understand its potential influence on the estimation of oscillatory parameters, we first established normative lifespan reference models for the aperiodic 1/f slope and offset. Using data from our large outpatient cohort, we modeled lifespan trajectories of the aperiodic slope and offset using scatterplot averaging, stratified by sex and the six major brain lobes.

Both aperiodic parameters showed generally monotonic age-related changes. Across the lifespan, the aperiodic slope progressively flattened: its typically negative exponent became less negative with increasing age, indicating a relative increase in higher-frequency background power (Fig. 6, 1/f Aperiodic Slope). In parallel, the aperiodic offset, which reflecting broadband power at the lowest frequency of interest (1 Hz), declined monotonically with age 6, 1/f Aperiodic Offset). These developmental and ageing patterns were similar across sexes and broadly conserved across cortical lobes, although the rates and magnitudes of change varied across developmental stages and regions. As in our oscillatory analyses, regional variability exceeded sex differences (Supplementary Fig. 4e,f).

**Figure 6:**
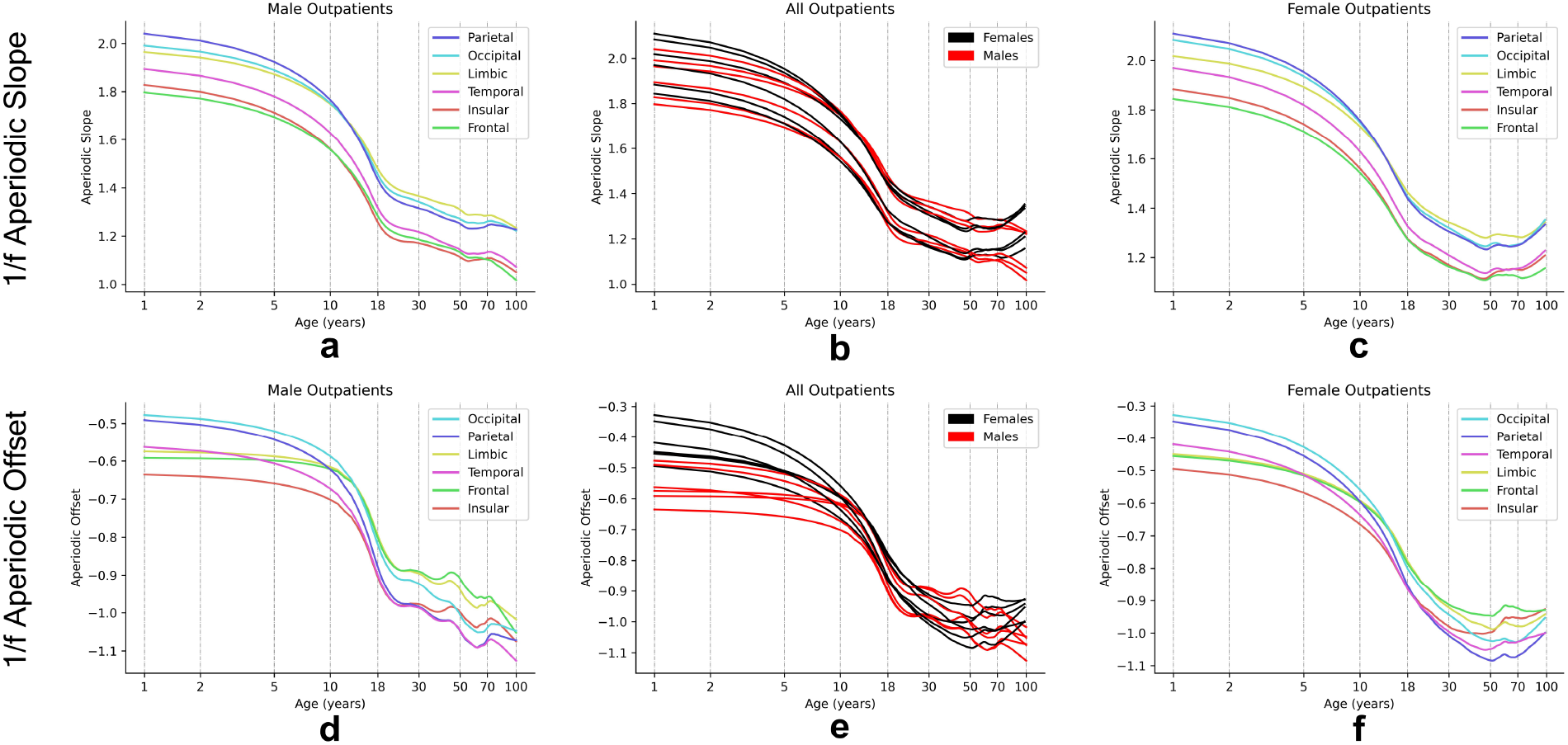
Changes in aperiodic 1/f activity parameters as a function of age across lifespan due to variation in two factors: brain region and sex. The age on the x-axis is represented using natural logarithmic scale. Both aperiodic slope and offset were characterized with progressively declining lifespan trajectories. This developmental trend across the lifespan was stable across sexes and six brain regions. **(a)** Lifespan reference charts for aperiodic slope for males. **(b)** Lifespan reference charts for aperiodic slope for males and females compared together visually. **(c)** Lifespan reference charts for aperiodic slope for females. **(d)** Lifespan reference charts for aperiodic offset for males. **(e)** Lifespan reference charts for aperiodic offset for males and females compared together visually. **(f)** Lifespan reference charts for aperiodic offset for females.

These normative charts therefore reveal a progressive flattening of the aperiodic slope and a concomitant reduction in aperiodic offset across the human lifespan, with consistent patterns across sexes and brain regions.

### Neurological and neuropsychiatric disorders show steeper aperiodic slopes and reduced offsets in EEG spectra

To elucidate the divergent behaviors of unadjusted and 1/f-adjusted alpha metrics across neurological and neuropsychiatric disorders, we next examined the underlying aperiodic 1/f activity. For each inpatient, aperiodic slope and offset were projected onto the sex- and lobe-specific normative lifespan charts, and age-corrected deviation scores were computed and summarized by diagnostic group, sex and cortical lobe. We then quantified the magnitude of these deviations using effect sizes (Fig. 7).

**Figure 7:**
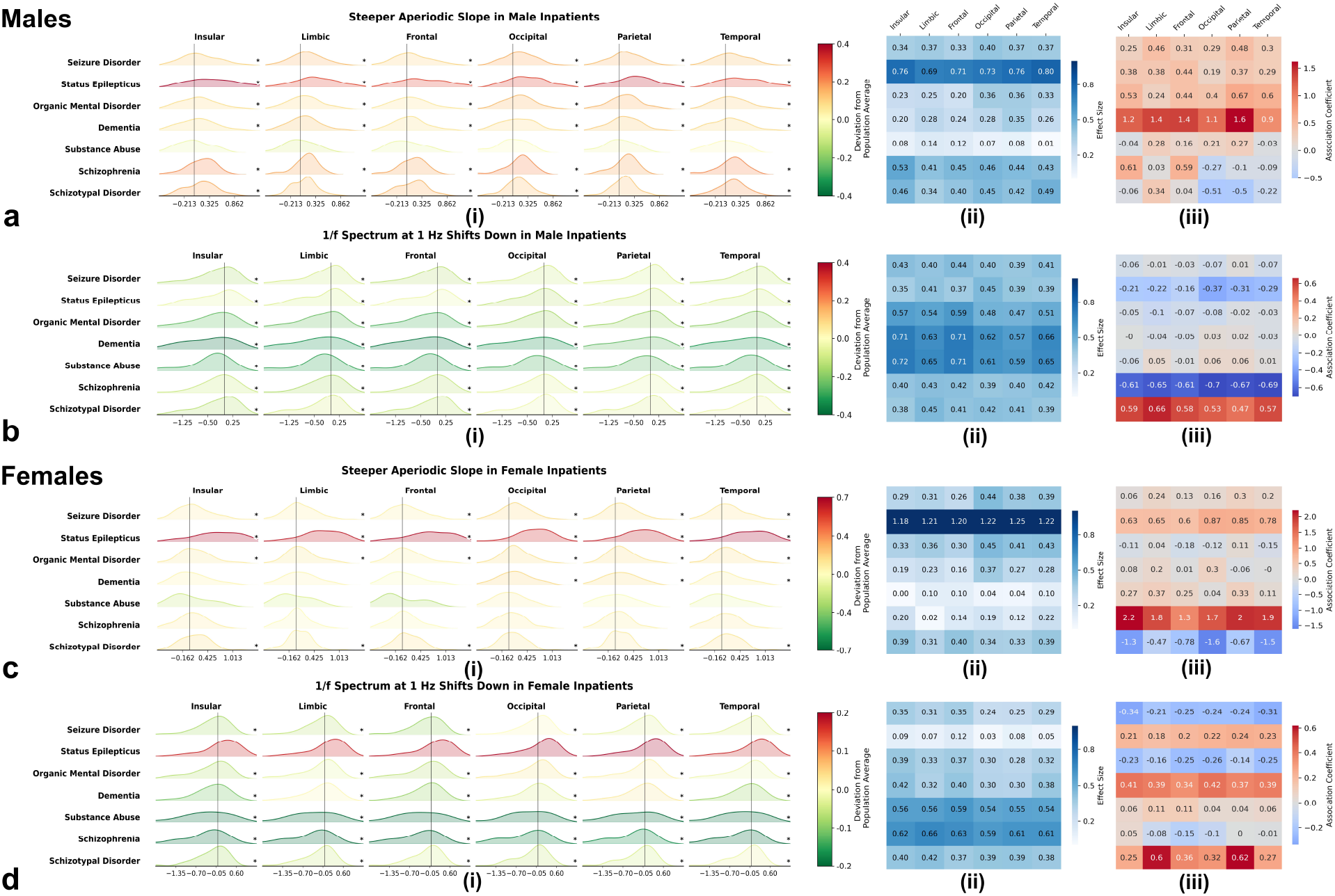
Aperiodic slope becomes steeper while aperiodic offset shifts down in neurological and neuropsychiatric disorders. Panels a-d profiles changes in aperiodic 1/f slope and offset in diagnostic groups relative to normative reference values, stratified by sex and brain regions. **a**,**c(i)** Deviations in aperiodic slope from normative reference charts. Red distributions show positive deviations indicating increased aperiodic slope (steeper slope) in wide range of disorders, with statistical significance (*p* < 0.1) marked by (*). This steepening was stable across brain regions but showed variability across sex and diagnostic groups. **a**,**c(ii)** Effect sizes of this steepening, with darker colors indicating stronger effects. The effect size varied across disorders. **b**,**d(i)** Deviations in aperiodic offset from lifespan reference charts. Green distributions represent negative deviations indicating reduced aperiodic offset (downward shift in power at low frequencies), with statistical significance (*p* < 0.1) marked by (*). This reduction was consistent across disorders, sexes, and brain regions. **b**,**d(ii)** Effect sizes of this reduction in offset. The strength of this effect varied across disorders. **a**,**b**,**c**,**d(iii)** Correlations between deviations in aperiodic slope/offset and comorbidity scores. No statistically significant correlations were observed.

The aperiodic slope was steeper in neurological and neuropsychiatric disorders relative to normative reference values, corresponding to more negative exponents in log–log power spectra (**Fig. 7 a,c(i)**). This steepening was broadly conserved across cortical regions, but showed diagnosis- and sex-specific exceptions: for example, it was absent in males with substance abuse disorders and in females with substance abuse or schizophrenia spectrum disorders. Effect sizes varied across diagnoses (**Fig. 7 a,c(ii)**), with the largest slope deviations observed in status epilepticus and the smallest in substance abuse.

Concurrently, the aperiodic offset exhibited a consistent downward shift, reflecting a broadband reduction in low-frequency power in the clinical groups, relative to the normative values (**Fig. 7 b,d(i)**). This reduction in offset was largely consistent across diagnostic groups, sexes, and brain regions, with a notable exception in female inpatients with status epilepticus who did not exhibit a clear offset shift. The magnitude of this downward offset shift varied by diagnostic groups (**Fig. 7 b,d(ii)**), being greatest in substance abuse and smallest in status epilepticus.

Overall, clinical groups showed consistent, coordinated changes in both aperiodic parameters: steeper slopes (more negative exponents) and reduced offsets.

### 1/f-adjusted alpha peak frequency is inversely related to 1/f-adjusted alpha power

To probe how changes in alpha-band timing and amplitude jointly manifest in neurological and neuropsychiatric disorders, we examined the relationship between deviations in alpha power and alpha peak frequency from their normative trajectories. For each inpatient EEG, we computed deviation scores (relative to age-, sex- and lobe-specific norms) for 1/f-adjusted alpha power and 1/f-adjusted APF. Then we quantified their correlation within diagnostic groups and cortical regions (Fig. 8).

**Figure 8:**
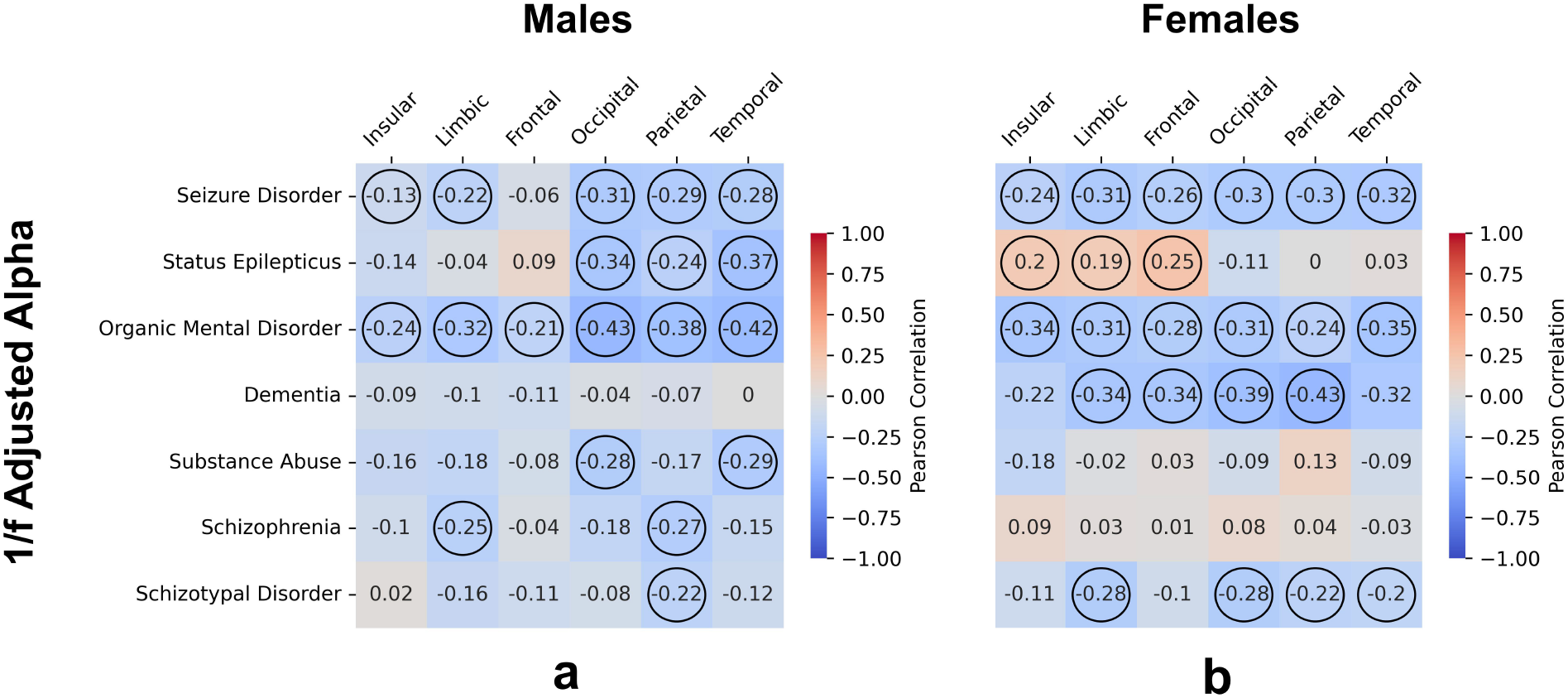
Inverse relation of 1/f-adjusted alpha frequency with 1/f-adjusted alpha power. Correlations between 1/f-adjusted alpha frequency and power, stratified by sex. Each cell in the matrix represents correlation value for a unique disorder-region pair. Positive correlations are presented in red and negative correlations in blue. Circles around the values indicate statistical significance. **a**,**b** Predominantly, significant negative correlations were observed (*p* < 0.1). This implied that a larger increase in adjusted alpha frequency resulted in smaller decrease in alpha power, bringing it closer to normative reference values.

Across disorders and lobes, deviations in 1/f-adjusted APF and 1/f-adjusted alpha power were generally negatively correlated. In other words, patients who showed greater acceleration of 1/f-adjusted APF (more positive deviation scores) tended to show weaker reductions in 1/f-adjusted alpha power (less negative deviation scores). This inverse relationship was broadly consistent across mental disorder groups and cortical regions, indicating that, once aperiodic activity is accounted for, faster alpha oscillations tend to co-occur with more “normal” alpha power in these clinical populations.

In contrast, correlations between deviations in absolute alpha power and unadjusted APF were more variable but predominantly positive (Supplementary Fig. 5). Thus, conventional alpha metrics suggest a different, and often opposite, coupling between frequency and power than that revealed by 1/f-adjusted measures, reinforcing the notion that aperiodic activity critically shapes apparent relationships among oscillatory parameters in neurological and neuropsychiatric disorders.

## Discussion

Our findings provide a unified, lifespan-based account of alpha-band and aperiodic EEG alterations in neurological and neuropsychiatric disorders. Using >22,000 outpatient EEGs, we derived sex- and lobe-specific normative “brain charts” for unadjusted and 1/f-adjusted alpha power and peak frequency, as well as aperiodic slope and offset, revealing conserved inverted-U trajectories for alpha power and frequency and progressive flattening and attenuation of aperiodic activity across the lifespan. Projecting inpatient data onto these norms, we found that neurological and neuropsychiatric disorders are broadly characterized by reduced alpha power, irrespective of whether spectra are 1/f-adjusted. But we also show a striking dissociation in alpha peak frequency: conventional unadjusted metrics suggest alpha slowing, whereas 1/f-adjusted estimates reveal alpha acceleration. We further show that steeper aperiodic slopes and reduced offsets accompany these paradoxical signatures. These changes produce a pronounced “spectral tilt” in neurological and neuropsychiatric disorders. When unadjusted spectra are analyzed using conventional methods, this tilt adds power at lower frequencies and reduces it at higher frequencies within the alpha-band range, mimicking a slowing of the alpha peak frequency and a loss of alpha power. Once the aperiodic component is explicitly parameterized and removed, the underlying alpha oscillations instead reveal an acceleration of peak frequency. This pattern provides a mechanistic explanation for the paradoxical contrast between unadjusted APF slowing and 1/f-adjusted APF acceleration observed in these clinical populations. Together, these results indicate that aperiodic changes are a core, lifespan-sensitive feature of brain dysfunction in mental illness and can fundamentally distort conventional interpretations of “alpha slowing.”

Beyond oscillatory measures, our results add nuance to ongoing debates about the role of aperiodic 1/f activity in neurological and neuropsychiatric disorders. We observed a consistent pattern of steeper aperiodic slopes (more negative exponents) and reduced offsets across diagnostic groups. The slope findings are broadly in line with prior reports of altered aperiodic exponents in psychiatric and neurological conditions [22–24, 26, 27], but the offset reductions diverge from some earlier studies that reported increased or unchanged offsets [22, 27, 28]. Our large, age-stratified sample and normative modeling framework suggest that these offset reductions are robust when age-related trajectories are explicitly accounted for. This raises the possibility that previous inconsistencies partly reflect smaller sample sizes, restricted age ranges, or inadequate age matching rather than true discrepancies in underlying physiology.

The pattern of steeper slopes in mental disorders is consistent with hypotheses linking aperiodic structure to excitatory–inhibitory (E/I) balance [17, 29]. Steeper slopes have been associated with relatively stronger inhibitory contributions to synaptic currents, whereas flatter slopes are linked to increased excitatory drive [14]. Many neurological and neuropsychiatric disorders are characterized by disrupted GABAergic inhibition and altered E/I balance [30, 31], which could simultaneously shape aperiodic background activity and the expression of rhythmic alpha oscillations [30, 32]. However, these associations remain indirect. Different disease stages, medications, and compensatory mechanisms could all modulate E/I balance in ways that do not map onto a simple “more inhibition indicates steeper slope” relation. Whether steeper slopes consistently reflect increased inhibition, or instead capture a broader class of pathophysiological changes, will require convergent evidence from pharmacological, invasive, and computational work.

While the pattern of steeper slopes in clinical populations is broadly consistent with the excitatory–inhibitory balance framework proposed by Gao et al. [29], in which steeper exponents reflect enhanced inhibitory synaptic contributions, recent biophysical modeling urges caution in interpretation. Brake et al. [33] demonstrated that aperiodic slope is shaped by multiple biophysical parameters beyond E/I ratio, including GABA receptor decay kinetics, membrane leak conductance, and synaptic time constants. These parameters can independently produce slope changes or even reverse the expected relationship between slope and E/I balance under certain membrane property configurations. Moreover, the foundational associations between E/I balance and spectral slope were established using local field potentials and electrocorticography, whereas scalp EEG introduces additional confounds. High-frequency neural activity attenuates disproportionately due to spatial averaging over small generator volumes [34], and EMG contamination above 20 Hz systematically flattens measured slopes [35], potentially creating pseudo-excitatory profiles in populations with altered muscle tone. Pharmacological studies further complicate interpretation. Propofol, for instance, steepens aperiodic slope primarily by prolonging GABA-A receptor open times rather than shifting network E/I balance [33].

Thus, while our observed steeper slopes and reduced offsets in clinical groups are consistent with altered inhibitory function, they likely reflect a composite signature of synaptic kinetic changes, membrane property alterations, and potentially measurement artifacts rather than a simple readout of cortical E/I balance. Nonetheless, these aperiodic features retain value as state- and disease-sensitive biomarkers, and their mechanistic role in producing spectral tilt remains robust regardless of their precise physiological origins. The spectral tilt we document systematically distorts conventional alpha metrics, creating apparent slowing that reverses to acceleration once aperiodic components are separated. This pattern holds independent of whether slope changes reflect true shifts in synaptic E/I balance, alterations in receptor kinetics, or confounds introduced by volume conduction and muscle artifacts. Future work combining scalp recordings with intracranial validation, computational modeling of source geometry effects, and pharmacological challenges with known E/I modulators will be essential to disambiguate the physiological contributors to aperiodic parameters in clinical populations.

An important conceptual caveat concerns the interpretation of the aperiodic component itself. The spectral parameterization approach we employed [14] treats the 1/f background as genuinely “aperiodic” activity, distinct from narrowband oscillatory peaks. In contrast, other neurophysiological models propose that 1/f-like spectra may emerge from the temporal integration of many partially overlapping oscillatory processes or network states across frequencies, rather than from a separate process [1]. On this view, what we treat as an aperiodic baseline could reflect the aggregate behavior of multiple, weakly synchronized rhythms rather than a distinct physiological substrate. The current data cannot adjudicate between these mechanistic accounts.

Despite this theoretical uncertainty, our results demonstrate the practical value of separating the 1/f component from narrowband alpha peaks. Regardless of whether 1/f activity reflects genuinely arrhythmic processes or emergent properties of complex oscillatory interactions, failing to model it explicitly leads to qualitatively different conclusions about alpha dynamics in neurological and neuropsychiatric disorders, most notably, an apparent “alpha slowing” that reverses to alpha acceleration after 1/f adjustment. Thus, even if the precise biophysical origin of the aperiodic term remains debated, its explicit treatment is essential to avoid conflating broadband spectral shifts with changes in alpha-band oscillations, particularly in lifespan- and disease-related contexts.

The emergence of alpha acceleration following 1/f adjustment in our transdiagnostic sample aligns with growing evidence that alpha oscillations can genuinely accelerate in specific neurological and neuropsychiatric populations. In post-traumatic stress disorder, combat veterans show markedly faster individual alpha peak frequency compared to trauma-exposed controls, alongside reduced alpha power and flatter aperiodic slopes [36], replicating earlier observations of elevated alpha frequency in PTSD [37]. Similarly, young children with autism spectrum disorder exhibit substantially higher peak alpha frequency than typically developing peers, a pattern Edgar et al. characterized as early maturation of alpha rhythms that normalizes across development as neurotypical children’s alpha frequencies progressively increase [38]. Yet the relationship between aperiodic activity and alpha parameters varies meaningfully across disorders. Chen et al. found that apparent alpha differences between ADHD and control children disappeared entirely when using aperiodic-corrected measures [39], suggesting that aperiodic changes rather than oscillatory abnormalities may be the primary spectral feature in some conditions. These contrasting patterns underscore that separating periodic from aperiodic components can reveal distinct pathophysiological signatures: genuine alpha acceleration in conditions like PTSD and autism that co-occur with flatter aperiodic slopes, versus predominantly aperiodic alterations in other disorders where conventional alpha metrics may misattribute broadband spectral changes to oscillatory dysfunction. Whether the co-occurrence of alpha acceleration and flatter slopes in hyperarousal-related conditions reflects shared biophysical mechanisms or independent pathophysiological processes remains an open question requiring convergent evidence from invasive recordings and computational modeling.

Beyond clarifying the aperiodic mechanism underlying the apparent “slowing” paradox, our findings point to an active, compensatory reconfiguration of alpha dynamics in diseased brains. After accounting for 1/f structure, neurological and neuropsychiatric disorders showed a coupled pattern: alpha power decreased, whereas alpha peak frequency increased, and these deviations were inversely related. Specifically, patients with larger frequency increases tended to show smaller reductions in alpha power. This inverse coupling was broadly conserved across diagnostic groups and cortical regions, suggesting that it reflects a generic network-level response rather than a disorder-specific signature.

This pattern is compatible with homeostatic plasticity mechanisms that act to stabilize network activity under chronic perturbation [40, 41]. Neurological and neuropsychiatric disorders and related brain insults are known to disrupt thalamocortical circuits that normally generate coherent alpha rhythms [42, 43], with reduced alpha power likely reflecting degraded large-scale synchrony as distributed networks lose the ability to sustain strong, spatially extended oscillations [9, 44]. Within this framework, an accompanying acceleration of alpha frequency could represent an emergent strategy to preserve effective communication and temporal coordination despite weakened synchrony. Computational models show that increased neuronal excitability can to some extent, compensate for reduced coupling by shifting networks into faster oscillatory regimes [45], and that systems near critical states maximize information transmission and dynamic range [46, 47]. The compensatory dynamics we observe, namely, lower alpha power but faster, more “tightly tuned” oscillations, may thus reflect attempts to retune damaged networks back toward a critical operating regime [48, 49].

These ideas are particularly salient in epilepsy, where hyperexcitability and impaired inhibitory control are defining features of the network [30, 50]. In such systems, faster background oscillations could arise as the network searches for a stable operating point under imbalanced excitation–inhibition, even as macroscopic synchrony (and thus alpha power) is compromised [51–53]. Whether these frequency accelerations are functionally beneficial, that is, supporting residual cognition and perception in the face of structural and neurochemical damage [54, 55] or instead reflect maladaptive plasticity that entrenches pathological states [56, 57] likely varies across disorders, stages and treatments. Longitudinal and interventional studies, combining 1/f-adjusted spectral metrics with behavioral and clinical outcomes, will be essential to determine when accelerated alpha represents an adaptive compensation and when it is a marker of failing homeostasis.

Our findings have immediate translational potential for clinical EEG interpretation. The normative reference charts we established enable quantitative, individualized assessment of alpha abnormalities relative to age-matched references, moving beyond qualitative descriptions of “slowing” or “abnormal” toward precise deviation scores. This approach offers several advantages: objective quantification of abnormality severity [58], the potential to detect subtle changes before overt symptoms [59], stratification of heterogeneous patient populations into neurophysiologically defined subgroups [60], and personalized monitoring of treatment response [61].

## Data and Code Availability

Data supporting the findings of this study include de-identified participant-level, lobe-wise averaged values for several spectral parameters analyzed in the manuscript. During peer review, these data and the associated code for the analyses will be made available to editors and reviewers upon request. After publication, access to the minimum dataset will be provided upon reasonable request to the corresponding author, subject to applicable ethics approvals and data-sharing agreements required for human participant data.

## Methods

### Subjects

We analyzed a large dataset of routine clinical electroencephalography (EEG) recordings collected between 2010 and 2018 from four public hospitals within the Fraser Health Authority in British Columbia, Canada. Patient demographic variables, including age and sex, were incorporated into our analysis alongside EEG recordings. EEG recordings were selected without bias, and males and females were analyzed separately. The Research Ethics Board at Simon Fraser University and Fraser Health Authority approved the ethics protocol on 1^*st*^ April 2022 (protocol number: H18-02728). Patients were broadly classified as outpatients or inpatients, following the categorization defined by the Canadian Institute for Health Information (CIHI). Our cohort represented a culturally and socioeconomically diverse population.

Outpatients were individuals who visited hospitals for check-ups but were not admitted. The outpatient group included a total of *N* = 22,094 patients, comprising 10,939 males (age: 43.34 ± 23.06 years) and 11,154 females (age: 44.05 ± 22.22 years). Inpatients were individuals who spent at least one night in the hospital and received acute care, excluding those only seen in emergency departments. Inpatients were categorized into Case Mix Groups (CMGs), defined by the CIHI based on International Statistical Classification of Diseases and Related Health Problems, 10th Revision, Canada (ICD-10-CA). This categorization included 21 CMG groups, with each group representing a Major Clinical Category (MCC). For this study, we selected MCC-17 (Mental Diseases and Disorders) as our inpatient category, as we wanted to focus on mental disorders and their impact on electrophysiological activity of the brain. Within MCC-17, we identified 7 inpatient sub-groups including seizure disorder, status epilepticus, organic mental disorder, dementia, substance abuse disorder, schizotypal disorder, and schizophrenia. The seven clinical groups were selected based on the criterion that the number of males or females in each group was at least greater than 50. The demographic characteristics of these inpatients are presented in Table 1. The inpatients were also classified according to five levels of comorbidity scores (0-4), which reflected overall health status. A score of 0 indicated good health, while a score of 4 signified poor health with an accumulation of multiple comorbidities.

### EEG Preprocessing

EEG recordings were acquired using standardized hardware and firmware across all sites. Each EEG station was equipped with a Natus Xltek EEG32U amplifier and gold-cup electrodes. A standard 10–20 electrode positioning system was used, collecting data from 20 electrode channels: FP1, FPZ, FP2, F3, F4, F7, F8, FZ, T3, T4, T5, T6, C3, C4, CZ, P3, P4, PZ, O1, and O2. The original sampling frequency for all EEG recordings was either 500 or 512 Hz.

Our EEG preprocessing pipeline included the following steps: First, EEGs were converted from the Native Natus proprietary format to the European Data Format (EDF). The EDF-formatted EEGs were then anonymized using PyEDFlib library[62] in Python. Next, a zero-phase, overlap-add finite impulse response band-pass filter with a Hamming window between frequency range 0.5-55 Hz was applied using the MNE-Python library [63]. This frequency range was selected to include all critical canonical brain rhythms while excluding noise above the 60 Hz powerline frequency. The EEGs were then resampled to 256 Hz for standardization. From the standardized EEG data, we removed flat intervals (digital zeros with minimum a peak-to-peak threshold of 1e-6), photic stimulation, and hyperventilation procedures. The final dataset consisted of 6-minute, 20-channel sensor space, Z-score normalized EEG recordings for each patient.

### Source Reconstruction and EEG Spectral Power Estimation

EEG source activity was reconstructed at 148 cortical regions of interest (ROIs) (74 per hemisphere) as defined in Destrieux atlas [25]. These ROIs were assigned to one of the six cortical regions: Insular, Limbic, Frontal, Occipital, Parietal, and Temporal lobes defined in the Destrieux atlas. Source reconstruction was performed using a beamforming approach [64]. First, forward solutions (lead fields) [65–67] were computed for each cortical source using MNE-Python library. This process employed a three-layer boundary element model (BEM) of the patient’s head, assigning standard conductivity values to the inner skull (0.3 S/m), outer skull (0.006 S/m), and skin (0.3 S/m). Next, neural dynamics at each cortical source were constructed using a scalar single-source minimum variance beamformer [64, 67]. The resulting time courses were normalized to account for depth-related biases. Specifically, we divided the time courses by the root mean square (RMS) amplitude of sensory noise projected by the beamformer to ensure uniform scaling. This normalization resulted in dimensionless pseudo-Z units, where pseudo-Z represents the total signal-to-noise (SNR) ratio (pseudo-Z = SNR + 1). Spectral Power was obtained for the EEGs in source space using Welch method [68] from Scipy library [69] in Python. The spectral power is estimated between frequency range of 1 Hz to 55 Hz, with a linear increment of 0.5 Hz. The final spectral power data for each EEG consisted of 148 ROIs source space, 109 linearly spaced frequency values.

### Alpha Extraction from EEG Spectral Power

From the source reconstructed EEG spectral power, we extracted alpha rhythm parameters (frequency and power) using two methods: (1.) Band pass filtering and (2.) Modeling EEG spectral power into periodic and aperiodic components. For band-pass filtering, we applied a band-pass filter to extract alpha frequency and power directly from EEG spectral power, defining alpha rhythms within the 8–13 Hz range. The frequency within this range with the highest power was selected as the dominant alpha frequency, referred to as ‘Unadjusted Alpha Frequency’. The corresponding power was defined as ‘Absolute Alpha Power’, with units of (pseudo-Z)2/Hz. This approach captured overall alpha activity within the EEG spectrum. For the second approach, we used an EEG spectral power modeling toolbox presented in [14]. This method decomposed EEG spectral power into aperiodic (1/f) component and a set of Gaussian distributions modeling spectral peaks associated with different oscillatory brain rhythms (alpha, beta, and others). The 1/f aperiodic component was characterized by its slope and offset. Aperiodic slope described the overall trend of the EEG spectral power. Offset determined the overall broadband shift of spectral power. The Gaussian peaks represented distinct oscillatory rhythms in the power spectrum. We defined the alpha rhythms within the 8-13 Hz range. If multiple peaks were detected within this range, one with the highest power was selected as the dominant alpha frequency, referred to as ‘Adjusted Alpha Frequency’, with corresponding power defined as ‘Adjusted Alpha Power’ (units: arbitrary units (*a*.*u*.)). This method isolated true alpha oscillations from EEG spectrum while accounting for aperiodic background activity.

Alpha frequency and power were computed individually for each ROI from both methods. We then performed region-wise averaging across ROIs to find a representative value of alpha frequency and power for the given lobe.

### Data Analysis and Statistics

Scatterplots were generated across lifespan for both aperiodic components and alpha rhythms in six brain regions, separately for male and female outpatients. To generate brain reference charts across lifespan from these scatterplots, we applied locally weighted scatterplot smoothing (LOWESS) method from Statsmodels [70] library. LOWESS is a non-parametric regression method that provides a smoothed fit to data through local polynomial fitting and is resistant to outliers.

To quantify regional brain heterogeneity and sex differences in lifespan trajectories, we computed pairwise differences between all six brain regions for each spectral parameter within individual participants. This yielded distributions of inter-regional differences across all possible comparisons (e.g., occipital minus temporal lobe values) across the entire cohort. Effect sizes (Cohen’s d) were calculated for these distributions of differences relative to zero difference. The effect sizes were summarized as a 6×6 heatmap for each parameter, where lower triangle represented effect sizes for differences in male lifespan trajectories across six brain regions; upper triangle represented effect sizes for differences in female lifespan trajectories across six brain regions; and the diagonal values represented differences between sexes in corresponding brain regions (Supplementary Fig. 4 **a**,**b**,**c**,**d**,**e**,**f**). This yielded a comprehensive map of regional brain variability that captured the magnitude of inter-regional and sex-related differences while controlling for individual-specific factors that affect absolute parameter values.

To quantify changes in oscillatory parameters across clinical groups, we employed a normative modeling approach by projecting inpatient alpha parameters onto the sex-specific outpatient reference charts described above. For each inpatient participant, we extracted the age-matched normative value from the appropriate LOWESS-fitted reference trajectory (male or female) at their specific age. We then calculated a deviation score by subtracting this age-matched reference value from the inpatient’s observed parameter value. Positive deviation scores indicate parameter values above the normative trajectory, while negative scores indicate values below the trajectory. This approach allowed us to quantify how individual inpatient measurements deviate from expected values based on the healthy outpatient reference population while accounting for age and sex. We then examined the distribution of these deviations across different brain regions and neurological and neuropsychiatric disorders, stratified by sex. The p-values for distribution of deviations were computed using 1-sampled t-test (compared with the population mean of 0). The effect sizes for these deviations were computed using Cohen’s d.

To interpret the magnitude of these effects, we applied conventional thresholds for Cohen’s d effect sizes, calculated as the mean deviation score divided by the standard deviation of deviation scores within each group. Effect sizes below 0.2 were considered small, indicating minimal deviation from the normative trajectory. Effect sizes between 0.2 and 0.5 were classified as medium, representing moderate deviations. Effect sizes between 0.5 and 0.8 were categorized as large, reflecting substantial deviations from age- and sex-matched reference values. Effect sizes greater than 0.8 were classified as very large, indicating pronounced differences between clinical groups and the reference outpatients’ trajectory. These standardized thresholds enabled consistent interpretation of deviation magnitudes across different brain regions and clinical populations.

To evaluate associations between deviations in alpha frequency and power and comorbidity scores, we applied the OrderedModel method from the Statsmodels library, which uses ordinal logistic regression approach for modeling correlations between an ordinal dependent variable (comorbidity levels in our case) and one or more independent variables (deviations in alpha rhythms parameters in our case). This model estimated odds ratios, indicating how deviations in alpha parameters influenced the likelihood of belonging to a higher comorbidity level. The method also returned a p-values corresponding to these associations.

Additionally, we computed Pearson correlations between the deviations in alpha frequency and power to assess the directional relationships between their deviations and their significance. All p-values were corrected for multiple comparisons using the Benjamini-Hochberg (BH) method with a family-wise error rate (*α*) of 10%, and the threshold for significance was kept at *p* < 0.1.

## Data Availability

All data produced in the present study are available upon reasonable request to the authors. The data will be provided as a lobe-averaged values.

